# Bi-directional effects between loneliness and substance use: Evidence from a Mendelian randomisation study

**DOI:** 10.1101/19006767

**Authors:** Harriet S R Greenstone, Robyn E Wootton, Abdel Abdellaoui, Damiaan Denys, Karin J H Verweij, Marcus R Munafò, Jorien L Treur

## Abstract

**Aims:** Loneliness and social isolation are associated with cigarette smoking and problematic alcohol use. Observational evidence suggests these associations arise because loneliness increases substance use, however there is potential for reverse causation (problematic drinking causing damage to social networks, leading to loneliness). With conventional epidemiological methods, controlling for (residual) confounding and reverse causality is difficult. In this study, we apply Mendelian randomisation (MR) to assess bi-directional causal effects between loneliness on the one hand and smoking behaviour and alcohol (ab)use on the other.

**Design:** We applied bi-directional MR using summary-level data of the largest available genome-wide association studies of loneliness (n=511,280), smoking (initiation (n=249,171), cigarettes-per-day (n=249,171) and cessation (n=143,852)), alcoholic drinks-per-week (n=226,223) and alcohol dependence (n=46,568), using independent samples. For each relationship, we selected genetic variants predictive of the exposure variable as instruments and tested their association with the outcome variable. Effect estimates for individual variants were combined with inverse-variance weighted regression (gene-outcome/gene-exposure association) and the robustness of these findings was assessed with five different sensitivity methods.

**Findings:** There was weak evidence of increased loneliness leading to higher likelihood of initiating smoking and smoking more cigarettes, and a lower likelihood of quitting smoking. Additionally, there was evidence that initiating smoking increases loneliness. We found no evidence of a causal effect between loneliness and alcohol (ab)use.

**Conclusions:** We report tentative evidence for causal, bidirectional, increasing effects between loneliness and cigarette smoking. These findings improve our understanding of the interrelatedness of smoking and loneliness, however, replication with better powered genetic instruments is recommended.

## Background

Extreme and prolonged loneliness is associated with worse physical and mental health^1,2^, with evidence that loneliness and social isolation are comparable in magnitude to other well-established risk factors for mortality^3^. One proposed explanation is that loneliness is associated with poor health behaviours^4^. Studies indicate 5-30% of adults are lonely^5,6,7,8^, suggesting high relevance for public health in identifying causal links between loneliness and health behaviours^6, 7^.

There is a particularly consistent link between loneliness and tobacco smoking and alcohol use; two of the most detrimental health behaviours worldwide. Lonely individuals are more likely to be cigarette smokers^4,9^, potentially resulting from smokers’ attempts to regain belonging in environments where smoking is socially acceptable^10^. Feelings of loneliness are associated with higher smoking in a nationally representative sample of adults^10^ and social support appears beneficial when considering and maintaining smoking cessation^11^. Similarly, greater daily alcohol use is associated with lack of social activity amongst older adults in the general population^12^ and clinical samples^2,13^.

Furthermore, there are strong genetic correlations between loneliness and increased alcohol dependence, smoking heaviness, likelihood to initiate smoking and decreased likelihood of smoking cessation^5^, suggesting possible causal pathways. However, in order to support a causal effect, we must first rule out residual confounding. For example, alcohol consumption is partly determined by societal attitudes to alcohol^14^ and stress (perhaps exacerbated by loneliness) may also play an indirect role in risky behaviours^15^ like excessive drinking. Furthermore, there remains potential for reverse causality; as problematic drinking may cause damage to and limit social networks, leading to loneliness.

To date, only observational studies have examined associations between substance use and loneliness. With observational data, it is difficult to control for the effects of residual confounding and reverse causation. In this study, we apply Mendelian randomisation (MR) to assess bi-directional causal effects between loneliness and smoking behaviour and loneliness and alcohol (ab)use.

## Methods

MR is an instrumental variable method, using genetic variants as a proxy for an exposure to estimate the effect of that exposure on an outcome^16^. MR can provide evidence of a causal effect that avoids bias from confounding and reverse causation, if the following hold: 1) genetic variants robustly predict the exposure, 2) genetic variants are not associated with confounders, and 3)genetic variants are only associated with the outcome through the exposure. The latter two assumptions can be violated by horizontal pleiotropy, which occurs when one genetic variant directly influences two traits, inducing spurious causal effect. We conduct multiple sensitivity analyses, each with different assumptions, to test for pleiotropy.

### Data

We applied bi-directional MR using summary-level data of published genome-wide association studies (GWAS) of loneliness (n=511,280)^5^, smoking (initiation (n=249,171), cigarettes-per-day (n=249,171 smokers) and cessation (n=143,852))^18^, and alcohol use (drinks-per-week (n=226,223)^17^ and alcohol dependence (n=46,568)^1^. The sample sizes for the smoking variables and for alcoholic drinks are considerably lower than in the original GWAS because we based our analyses on summary-level data with UKBiobank and 23andMe, Inc. samples removed. This was to avoid sample overlap which can cause bias towards the observational association.

### Statistical analyses

Analyses were conducted using the TwoSampleMR package for R^19, 20^. Briefly, independent variants that passed the genome-wide level of significance (p<5×10^−8^) in the exposure GWAS were selected as instruments. This provided 16 single nucleotide polymorphisms (SNPs) for loneliness^5^, 378 SNPs for smoking initiation^17^, 99 SNPs for drinks-per-week ^17^ and 11 SNPs for alcohol dependence^18^. Because there were relatively few genome-wide significant SNPs for loneliness and alcohol dependence, we added instruments with relaxed *p*-value thresholds of p<1×10^−5^ for both. SNPs were clumped for independence at *r*^2^ <0.01 and 10,000 kb^19^. Next, these sets of SNPs were identified in the outcome GWAS. Cigarettes-per-day and smoking cessation could only be used as outcome variables because those GWAS only contained ‘ever-smokers’; and there was insufficient information to stratify by smoking status in the loneliness GWAS.

The main analysis was an inverse-variance weighted (IVW) regression model (SNP–outcome association/SNP–exposure, whereby each SNP is weighted according to the inverse of its variance). We applied five sensitivity methods; weighted median^21^, weighted mode^22^, MREgger^23^, Steiger filtering^24^, and generalized summary-based MR(GSMR)^25^. A consistent result across these methods would provide the greatest confidence a causal effect. The reliability of MR Egger is evaluated using the I^2^ GX statistic^26^. We also calculated the mean F-statistic to test instrument strength (F>10 being sufficiently strong) and Cochran’s Q to estimate heterogeneity between the SNP effects which might suggest pleiotropy.

## Results

### Causal effects of loneliness on substance use

With the *p*<1×10^−5^ threshold only, there was weak evidence of increased loneliness leading to a higher likelihood of initiating smoking (IVW *β*=0.10, 95% CI=0.06 to 0.13, *p*=4.6e-05, see Table 1) and smoking more cigarettes-per-day once started (IVW *β*=0.09, 95% CI=0.03 to 0.15, *p*=0.005). With both *p*-value thresholds, there was weak evidence for increased loneliness decreasing the odds of smoking cessation (IVW (*p*<5e-08) *β*=-0.09, 95% CI=-0.19 to 0.01, *p*=0.075; IVW (*p*<1e-05) *β*=-0.09, 95% CI =-0.13 to −0.05, *p*=1.3e-04). Mostly, results were consistent across the weighted median and GSMR sensitivity methods in effect size and direction of effect (with slightly weaker statistical evidence), but not with the weighted mode. MR-Egger results were not reported due to low reliability based on the I^2^ GX (Supplementary Table 3). For loneliness-smoking initiation and loneliness-cigarettes-per-day there was evidence of heterogeneity as measured with Cochran’s Q, while for loneliness-smoking cessation that was not the case (Supplementary Table2). Steiger filtering showed that all (except one) SNPs explained more variance in the exposure than in the outcomes, suggesting the effects were not due to reverse causation (Supplementary Table5). We found no clear evidence for causal effects of loneliness on alcohol (ab)use.

**Table 1.** Results of the two sample, bidirectional Mendelian randomization from loneliness to substance use and from substance use to loneliness

| Exposure | Outcome | *n* | IVW |  | Weighted median |  | Weighted mode |  | MR-Egger |  | *n* | GSMR |  |
| --- | --- | --- | --- | --- | --- | --- | --- | --- | --- | --- | --- | --- | --- |
|  |  | SNPs | beta (95% CI) | *p* | beta (95% CI) | *p* | beta (95% CI) | *p* | beta (95% CI) | *p* |  | beta (95% CI) | *p* |
| Loneliness *p*<5e-08 | Smoking initiation | 14 | 0.03 (-0.09 to 0.15) | 0.574 | 0.08 (-0.02 to 0.18) | 0.133 | 0.10 (-0.08 to 0.27) | 0.284 | *n.a.* | *n.a.* | 12 | 0.05 (-0.03 to 0.13) | 0.191 |
| Loneliness *p*<1e-05 | Smoking initiation | 108 | 0.10 (0.06 to 0.13) | 4.6e-05 | 0.08 (0.02 to 0.13) | 0.003 | 0.06 (-0.06 to 0.18) | 0.339 | *n.a.* | *n.a.* | 97 | 0.10 (0.06 to 0.14) | 4.0e-08 |
| Loneliness *p*<5e-08 | Cigarettes-per-day | 13 | 0.09 (-0.09 to 0.27) | 0.312 | 0.00 (-0.17 to 0.18) | 0.970 | -0.02 (-0.28 to 0.24) | 0.883 | *n.a.* | *n.a.* | 13 | 0.08 (-0.04 to 0.19) | 0.222 |
| Loneliness *p*<1e-05 | Cigarettes-per-day | 105 | 0.09 (0.03 to 0.15) | 0.005 | 0.02 (-0.06 to 0.10) | 0.623 | -0.03 (-0.23 to 0.17) | 0.804 | *n.a.* | *n.a.* | 98 | 0.09 (0.03 to 0.14) | 0.002 |
| Loneliness *p*<5e-08 | Smoking cessation | 13 | -0.09 (-0.19 to 0.01) | 0.075 | -0.06 (-0.20 to 0.08) | 0.374 | 0.04 (-0.19 to 0.28) | 0.715 | *n.a.* | *n.a.* | 13 | 0.08 (-0.02 to 0.18) | 0.099 |
| Loneliness *p*<1e-05 | Smoking cessation | 100 | -0.09 (-0.13 to -0.05) | 1.3e-04 | -0.06 (-0.12 to 0.00) | 0.060 | -0.02 (-0.18 to 0.13) | 0.750 | *n.a.* | *n.a.* | 95 | 0.08 (0.04 to 0.12) | 0.001 |
| Loneliness *p*<5e-08 | Drinks-per-week | 14 | 0.01 (-0.11 to 0.13) | 0.865 | 0.01 (-0.12 to 0.15) | 0.801 | 0.04 (-0.15 to 0.24) | 0.690 | *n.a.* | *n.a.* | 13 | -0.02 (-0.11 to 0.08) | 0.729 |
| Loneliness *p*<1e-05 | Drinks-per-week | 107 | -0.03 (-0.07 to 0.01) | 0.269 | -0.03 (-0.09 to 0.03) | 0.359 | -0.03 (-0.19 to 0.13) | 0.703 | *n.a.* | *n.a.* | 99 | -0.03 (-0.07 to 0.01) | 0.189 |
| Loneliness *p*<5e-08 | Alcohol dep | 16 | 0.09 (-0.19 to 0.36) | 0.533 | 0.26 (-0.09 to 0.61) | 0.140 | 0.34 (-0.23 to 0.91) | 0.259 | *n.a.* | *n.a.* | 15 | 0.07 (-0.20 to 0.34) | 0.602 |
| Loneliness *p*<1e-05 | Alcohol dep | 120 | 0.10 (-0.02 to 0.22) | 0.114 | 0.16 (-0.01 to 0.34) | 0.060 | 0.19 (-0.24 to 0.62) | 0.399 | *n.a.* | *n.a.* | 110 | 0.11 (-0.01 to 0.23) | 0.070 |
| Smoking initiation | Loneliness | 287 | 0.30 (0.22 to 0.38) | 2.8e-13 | 0.22 (0.12 to 0.31) | 2.2e-06 | 0.21 (0.03 to 0.38) | 0.022 | *n.a.* | *n.a.* | 255 | 0.23 (0.17 to 0.29) | 1.1e-14 |
| Drinks-per-week | Loneliness | 68 | 0.09 (-0.02 to 0.22) | 0.076 | 0.07 (-0.05 to 0.19) | 0.286 | 0.08 (-0.10 to 0.26) | 0.357 | 0.00 (-0.31 to 0.32) | 0.992 | 56 | 0.04 (-0.04 to 0.12) | 0.289 |
| Alcohol dep *p*<5e-08 | Loneliness | 6 | 0.06 (-0.02 to 0.13) | 0.162 | 0.06 (-0.02 to 0.14) | 0.082 | 0.03 (-0.07 to 0.13) | 0.578 | 0.11 (-0.32 to 0.54) | 0.644 | 6 | 0.05 (-0.03 to 0.13) | 0.139 |
| Alcohol dep *p*<1e-05 | Loneliness | 16 | 0.00 (-0.04 to 0.04) | 0.911 | 0.00 (-0.06 to 0.06) | 0.865 | -0.01 (-0.08 to 0.07) | 0.880 | -0.06 (-0.25 to 0.14) | 0.576 | 16 | 0.00 (-0.04 to 0.04) | 0.972 |

IVW = Inverse Variance Weighted regression, GSMR = Generalized Summary-based MR. *n.a.*: MR-Egger results not reported because of limited reliability based on the I-squared measure being <0.60.

### Causal effects of substance use on loneliness

Across the different MR methods, there was consistent evidence of a causal influence of smoking initiation on increased loneliness (IVW *β*=0.30, 95% CI=0.22 to 0.38, *p*=2.8e-13), despite the instrument being relatively weak (F-statistic Supplementary Table1). However, there was particularly strong evidence of SNP-heterogeneity (Cochran’s Q 729.30, *p*=1.5e-40), implying there could still be some horizontal pleiotropy for this relationship. With Steiger filtering the majority of SNPs, 277 of 287, explained more variance in the exposure than the outcome. There was very weak evidence of an increasing effect of drinks-per-week on loneliness with IVW (*β*=0.09, 95% CI=-0.02 to 0.22, *p*=0.076), but this did not hold up with any of the sensitivity methods. Finally, there was no clear evidence for causal effects of alcohol dependence on loneliness.

## Discussion

This is the first MR study exploring bi-directional associations between loneliness and substance use. We report tentative evidence for bidirectional effects between loneliness and smoking behaviour, such that loneliness increases the odds of initiating smoking, heavier smoking once started, and finding it difficult to quit, and that smoking initiation increases the odds of experiencing loneliness.

The fact that our evidence was not consistent for all sensitivity methods could be due to limited power and warrants further replication when larger sample sizes are available. Our findings that loneliness increases smoking are in line with pre-existing observations that lack of social connectedness may lead to increased smoking and difficulty in quitting^27,28^. Our finding of potential causal effects of smoking on loneliness is particularly interesting, and consistent with recent results from an MR study that found that smoking increases depressive symptoms^29^. The mechanism for this may result from inhaled nicotine acting on nicotinic cholinergic receptors, disrupting the release of neurotransmitters such as dopamine and serotonin, well-established players in the aetiology of depression. Feelings of loneliness and depressive symptoms are highly phenotypically and genetically correlated^30, 5^ and so it seems that the (biological) effects of smoking that could lead to depressive feelings plausibly also lead to higher odds of experiencing loneliness. Other constituents of tobacco smoke could also impact neurotransmitters, with suggestions that MAO inhibition is also implicated^30^.

Apart from some weak evidence that having more alcoholic drinks-per-week increases loneliness, which was not supported by any of the sensitivity methods, there was no clear indication of causal effects between loneliness and alcohol use. Further studies with better-powered genetic instruments are needed to fully assess the link between drinks-per-week and loneliness. Future work should also look at drinking frequency as there may be complexities such that loneliness is associated with extremes of drinking frequency rather than moderate drinking^31^. There may also be differences when considering frequency compared with quantity of alcohol consumption per occasion, with evidence indicating the former is generally positively correlated with health outcomes, whilst the latter is negatively correlated^32^. In addition, we found no clear evidence overall for effects between loneliness and alcohol dependence. While this could be due to low statistical power, it does align with some literature showing no evidence of an association between loneliness and at-risk drinking, binge drinking and extreme alcohol use^31 33^.

There are some important strengths to our study. We are first to apply MR using the largest available GWAS to examine bi-directional results between smoking and loneliness and alcohol and loneliness. We maximised the robustness of our findings by using a wide range of MR (sensitivity) methods, attempting to overcome the issue of horizontal pleiotropy. Applying multiple different MR methods, which each make different assumptions about the nature of pleiotropy, aims to overcome any individual limitation of a specific method. As required for MR, we also excluded overlapping samples; for example, if the GWAS for the exposure had contained the same people as for the outcome, then this result would be biased towards the observed estimate^16^.

However, there are some limitations. First, the genetic instrument for loneliness was relatively weak due to the small number of genome-wide significant SNPs. Therefore, we relaxed *p*-value thresholds for instrument selection to increase the number of SNPs in the instrument. This could increase the likelihood of pleiotropy, which we attempted to overcome by using a variety of sensitivity methods. While the instrument for smoking initiation was also of arguably low strength (given the F-statistic <10), we did find considerable evidence for causal effects. While it therefore did not appear to have limited our findings, replication of these results with stronger genetic instruments is advised. The loneliness GWAS is predominately based on the UKBiobank cohort; even after controlling for population structure, coincident structure and geographic clustering remain^34,35^, potentially introducing bias. We attempted to overcome this by ensuring the outcome sample did not overlap with UKBiobank. Additionally, there may be selection bias; UKBiobank participants are well-educated, healthier and less likely smokers compared to the general population^36^. Loneliness rates may therefore not be representative. If smoking, alcohol use, and/or loneliness reduce likelihood of participating in the UKBiobank, we would lack results for those most significantly affected – meaning our results may underestimate the association. Finally, our judgement of loneliness as a nominal variable is arguably flawed; failing to account for those intermittently but intensely lonely, or those with limited social connectedness, but who enjoy or benefit from this solitude^33,37^.

## Conclusions

In conclusion, we are first to examine bi-directional effects between loneliness and health behaviours with an MR framework. Although there was no clear evidence for effects between loneliness and alcohol (neither drinks-per-week nor alcohol dependence), there was moderate evidence for bi-directional effects between loneliness and smoking, which is supported by existing literature. We recommend our analyses be repeated using a stronger genetic instrument for loneliness in the future, which would increase the power of these findings. For now however, our findings are of relevance for population health. The negative health impacts of both smoking and loneliness have been established and addressing these factors in conjunction with a newfound understanding of their interrelatedness, seems an important public health goal. This could include an increased emphasis on social and interpersonal methods for smoking cessation and a greater recognition of the impact of loneliness on individuals using existing smoking cessation services.

## Data Availability

This paper uses summary statistics from publicly available genome-wide association studies (GWAS).

## Acknowledgements

MRM is a member of the UK Centre for Tobacco and Alcohol Studies, a UKCRC Public Health Research: Centre of Excellence. Funding from British Heart Foundation, Cancer Research UK, Economic and Social Research Council, Medical Research Council, and National Institute for Health Research, under the auspices of the UK Clinical Research Collaboration, is gratefully acknowledged. This work was supported by the Medical Research Council Integrative Epidemiology Unit at the University of Bristol, which is supported by the Medical Research Council and the University of Bristol (grant MC_UU_12013/7). JLT is supported by a Rubicon grant from the Netherlands Organization for Scientific Research (NWO; grant number 446-16-009) as well as a Veni grant (NWO; grant number 016.Veni.195.016). KJHV, AA, and JLT are supported by the Foundation Volksbond Rotterdam. AA is supported by ZonMw grant 849200011 from The Netherlands Organisation for Health Research and Development. This study was supported by the NIHR Biomedical Research Centre at the University Hospitals Bristol NHS Foundation Trust and the University of Bristol. The views expressed in this publication are those of the authors and not necessarily those of the NHS, the National Institute for Health Research or the Department of Health and Social Care.

